# Tobacco 21: Retailer Perceptions, Experiences, and Challenges Before and After a New Minimum Legal Sales Age Law in Maryland

**DOI:** 10.1101/2021.09.03.21250481

**Authors:** Candace Walsh, Alanna Biblow, John Charles, Patrick Madden, Dawn Berkowitz, Nikardi Jallah, Dana Moncrief

## Abstract

**Objectives:** By raising the legal age of sale from 18 to 21 years old, Tobacco 21 laws (T21) are reshaping the tobacco landscape in the United States. In 2019, Maryland became the 13th state to adopt T21. This study is among the first to examine the perception, awareness, and ease of compliance of T21 among tobacco retailers through a two-wave survey conducted pre-T21 and post-T21.

**Design:** Surveys were conducted among the state’s more than 6,000 registered retailers prior to the state’s T21 law going into effect and eight months later after T21 had been enacted. The pre-T21 and post-T21 survey instruments measured retailer awareness, support, and perceived ability to comply with T21, current tobacco practices, revenue from tobacco products, and challenges faced in compliance. 414 retailers completed the pre-T21 survey and 360 completed the post-T21 survey. The final survey data was weighted to reflect the geographic distributions of licensed tobacco retailers in Maryland.

**Results:** There was no significant difference in retailer awareness, support and challenges comparing pre and post T21. One notable caveat is lack of consumer knowledge about T21, resulting in retailers being responsible for educating customers.

**Conclusions:** The evaluation of T21 and impact to retailers is a valuable tool in determining the law’s success and barriers to implementation. Results support the idea that T21 laws have had a minor impact on how retailers conduct their business. As a result of the communication and materials provided by the state, retailers largely understand the law and how to adhere to guidelines.

## Introduction

On October 1, 2019, Maryland became the 13^th^ state in the United States to adopt a “Tobacco 21” or “T21” law, raising the minimum legal sales age (MLSA) for tobacco products from 18 to 21.^i^ In addition to raising the legal age, there were several changes to strengthen the existing law, including altering the term Electronic Nicotine Delivery Systems (ENDS) to Electronic Smoking Devices (ESD). The term ESD not only includes items such as e-cigarettes, vapes, pod devices such as Juul, electronic cigars, as well as any component or accessory of such a device but also encompasses a broader range of products. This term ensures that all pre-filled vaping devices and e-liquids or “juice,” as well as future products, are covered by the law, even if they claim not to contain nicotine. The new law also defined ESDs as a tobacco product and removed possess, use, and purchase (aka “PUP”) laws that cited the users. Removal of the PUP laws, places responsibility on the adult retailer instead of underage individuals obtaining the products. The law also included an exemption for active duty military personnel under 21 who had a valid military ID and required tobacco retailers to display appropriate signage.^ii iii^

Maryland and like-minded states have been adopting laws that raise the MLSA of tobacco products to 21 with the intent to reduce smoking rates and prevent the initiation of tobacco use among youth and young adults. In 2018, cigarette use among youth in Maryland continued to trend downward, with five percent of high school-age youth reporting current (past 30 day) use of these products. Use of ESDs remains concerning, with approximately 40% of Maryland high school students reporting having ever tried an ESD and 23% reporting current use of these products.^iv^

According to the U.S. Centers for Disease Control and Prevention (CDC), tobacco use and addiction are usually established during adolescence, with nine out of ten cigarette smokers trying their first cigarette by the age of 18.^v^ Findings from the 2018 Maryland Behavioral Risk Factor Surveillance System (BRFSS) indicate that initiation of cigarette smoking predominantly begins before the age of 21 and that among current smokers, 84% reported smoking their first cigarettes prior to age 21.^vi^ The retail environment remains a source of access and exposure to tobacco products for those under 21. Among Maryland teens who use ESDs, nearly 10% reported that they purchased them from a store.^vii^ However, similar to cigarettes, the usual source of ESDs differed by age at access, with high school youth aged 18 and older making direct purchases at a store or gas station (51.5%) or borrowing (18.9%) while high school youth under age 18 borrowed (44.5%) or gave someone else money to make the purchase for them (19.6%). Retailer compliance is vital to the success of T21. T21 policies seek to reduce tobacco use and initiation among youth and young adults by limiting underage access to tobacco products in stores and among peers, while changing the social norms for tobacco use among adolescents.^viii ix x^

In coordination with local health departments and statewide partners, the Maryland Department of Health (MDH) provided information to retailers to assist retailers in implementing this change effectively. In preparation for Maryland’s T21 law, MDH took several key steps to increase awareness about the new law to retailers and residents, including issuing three press releases, updating the Responsible Tobacco Retailer website (NoTobaccoSalesToMinors.com) and conducting several interviews. Additionally, MDH updated its Responsible Tobacco Retailer Media Campaign with a new message, “21 or None”. This campaign creative was used to update retailer materials, advertisements, and the website www.NoTobaccoSalesToMinors.com.

Following Maryland’s T21 implementation, on December 20, 2019 the United States government enacted legislation that raised the national MLSA of tobacco from 18 to 21. This federal legislation broadened T21 from nineteen states and Washington, D.C. who had already passed Tobacco 21 laws, to cover the entire U.S. Despite state and federal movement toward T21, studies evaluating the impact of T21 laws on the retail environment are limited. California conducted a retailer poll seven months following the implementation of their T21 law in 2016 and Massachusetts conducted a study of retailers in 2019 after T21 was in effect in 164 local communities.^xi xii^ Our study sought to add to this literature and understand retailer readiness, awareness, and compliance with the Maryland T21 law. Two waves of the survey were completed to understand retailer perceptions and practices: before and after T21 took effect in Maryland.

The pre-T21 survey was conducted between September 16, 2019 and September 30, 2019, just prior to the T21 law going into effect. The post-T21 survey was planned to take place at least sixth months after implementation. Despite the COVID-19 pandemic, post-T21 was conducted between May 17, 2020 and June 20, 2020, as retailers navigated operating their business under COVID-19 guidelines.

## Methods

To examine the perception, awareness, and expected ease of compliance of T21 among tobacco retailers in Maryland, surveys were conducted prior to the state’s T21 law going into effect and eight months later after T21 had been enacted.

The pre-T21 and post-T21 survey instruments measured retailer awareness, support, and perceived ability to comply with T21; current tobacco practices; revenue from tobacco products; challenges faced and opportunities to better support retailers in complying with T21 and other tobacco sales laws. The surveys received approval from the MDH Institutional Review Board (IRB) for the Protection of Human Subjects in Research.

### Sample

The target population for the research was all licensed tobacco retailers in Maryland, provided through a master list MDH obtained from the Maryland Office of the Comptroller. This list consisted of over 6,000 retailers of various types from across the state and included mailing addresses for all retailers. A limited number of email addresses were available in the list. Reverse lookups were used to identify telephone numbers for retailers when available based on their address. A census was conducted among all retailers for both the pre-T21 and post-T21 surveys. The research goal was to complete a total of 400 surveys for each survey wave, to provide a margin of error of ±5% at 95% confidence.

### Data Collection

A total of 414 retailers participated in the pre-T21 survey which took place from September 16, 2019 through September 30, 2019 and 360 participated in the post-T21 survey, which was conducted from May 22, 2020 to June 21, 2020. Data collection used a multi-mode approach that included an initial push-to-web and/or letter/email that invited retailers to participate in an online survey. Following the initial letter/email, those not responding to the survey were called on the phone and were given the opportunity to complete the survey via telephone or reminded to complete it online. The overall response rate for the pre-T21 survey was 11% and the post-T21 survey was six percent.

The surveys were directed to the owner, manager, or clerk at the store with sufficient knowledge about store tobacco sales policies and procedures. Written consent was obtained before starting the online survey. Respondents were screened to ensure that they were eligible to complete the survey (that is, they worked at a store that sold tobacco products). Participation in the survey was voluntary, and respondents were able to withdraw at any time during the survey or skip any question that they were unable or unwilling to answer. The surveys consisted of both multiple-choice and open-ended questions and took an average of 16 minutes to complete.

### Weighting

The final survey data were weighted to reflect the geographic distributions of licensed tobacco retailers in Maryland. Design weights were calculated separately for each of six (6) Maryland regions (Baltimore City, Capital, Central, Eastern Shore, Southern, Western). These designs weights were equal to the inverse of the probability of selecting a retailer within each region (i), or:

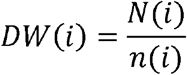

The size of the final weighted data for the pre-T21 survey was 6,091 and 6,251 for the post-survey, reflecting the total number of licensed tobacco retailers in Maryland according to the master list at the time of each survey.

### Statistical Analysis

All statistical analyses were performed using IBM SPSS Statistics 24 software. Analysis was conducted on the entire sample and comparisons were made between certain subgroups of interest, including store type, region, and respondent role at the store. Confidence intervals were calculated to determine whether there were significant differences between survey results when comparing all survey respondents’ characteristics. Statistically significant differences are noted where the 95% confidence intervals for the given point estimates do not overlap.

## Results

The results presented in this section show the differences in retailer perception and awareness between the pre-T21 survey and post-T21 survey. Table 1 shows the sample characteristics, including region, store type and respondent role. Respondents most often worked in a convenience store with gas, comprising of 26% of the pre-T21 respondents and 30% of the post-T21 respondents, followed by liquor stores (23%, 23%), and a convenience store without gas (16%, 20%). Those working for mass merchandisers made up the smallest group at 2% pre-T21 and 1% post-T21. Store owners were the most likely to have completed the surveys; 41% of respondents were owners pre-T21 and 56% post -T21, followed by managers (48%, 39%) and clerks (7%, 4%). The composition of respondents did not change significantly between the pre-T21 and post-T21 survey. The respondents were distributed evenly across regions.

**Table 1:** Population and Sample Characteristics.

|  | **Pre-T21** |  | **Post-T21** |  |
| --- | --- | --- | --- | --- |
|  | **Population**<br>**N= 6,091** | **Sample**<br>**n=414** | **Population**<br>**N=6,251** | **Sample**<br>**n=360** |
| **Region** |  |  |  |  |
| Baltimore City | 21% | 18% (72) | 27% | 24% (70) |
| Capital | 27% | 33%<br>(135) | 28% | 29% (87) |
| Central | 32% | 31%<br>(125) | 25% | 24% (77) |
| Eastern Shore | 10% | 10% (41) | 10% | 14% (44) |
| Southern | 6% | 5% (21) | 5% | 6% (22) |
| Western | 5% | 4% (15) | 4% | 4% (12) |
| **Store Type** |  |  |  |  |
| Convenience store<br>(without gas) | NA | 16% (63) | NA | 20% (63) |
| Convenience store<br>(with gas) | NA | 26%<br>(100) | NA | 30% (94) |
| Liquor store | NA | 23% (92) | NA | 23% (73) |
| Grocery store | NA | 14% (51) | NA | 11% (35) |
| Mass merchandisers | NA | 2% (8) | NA | 1% (2) |
| Tobacco shop | NA | 3% (22) | NA | 5% (15) |
| Vape shop | NA | 3% (28) | NA | 1% (4) |
| Something else (bars,<br>restaurants, head shop,<br>mechanic, golf course) | NA | 9% (33) | NA | 8% (24) |
| Pharmacy | NA | 3% (12) | NA | - |
| None of the above | NA | 2% (4) | NA | 1% (2) |
| **Respondent** |  |  |  |  |
| Owner | NA | 41%<br>(167) | NA | 56% (173) |
| Manager | NA | 48%<br>(197) | NA | 39% (121) |
| Clerk | NA | 7% (29) | NA | 4% (12) |
| Something else | NA | 4% (16) | NA | 2% (5) |

*Note: Percentages of sample are unweighted. NA indicates data is not available.*

Table 2 displays results for awareness, support and expected impact from the T21 law. Results between the pre-T21 and post-T21 survey showed little difference. Awareness of T21 was high among all respondents in both waves of the survey (97% pre-T21, 96% post T21), and nearly three-quarters supported the change in the law. Over two-thirds of pre (67%) and post (68%) survey respondents felt the increased sales age would or had caused them to card/ID customers purchasing tobacco products more often. While some retailers had concerns about the impact of T21 on their business operations, only 19% pre-T21 said they were significantly concerned, which declined slightly to 15% post-T21.

**Table 2:** Awareness, Support, and Impact of T21.

|  | **Pre-T21**<br>**n=412** |  | **Post-T21**<br>**n=358** |  |
| --- | --- | --- | --- | --- |
|  | **Yes (%)** | **No (%)** | **Yes (%)** | **No (%)** |
| Awareness of T21 law in Maryland | 96.8 (401) | 2.1 (7) | 98.5 (353) | 0.6 (2) |
| Aware of “21 or None” materials from the MDH for retailers (including signs, posters, window clings) | - | - | 80.9 (272) | 12.1 (41) |
| Support for T21 in Maryland | 71.6 (288) | 15.0 (69) | 69.6 (245) | 12.6 (43) |
| Card or check IDs of customers trying to buy tobacco more often because of T21 (% Yes) | 67.2 (279) | 30.7 (124) | 68.0 (229) | 30.5 (104) |
|  | **Easy (%)** | **Difficult (%)** | **Easy (%)** | **Difficult (%)** |
| Ease of compliance with T21 | 63.3 (265) | 4.3 (19) | 63.9 (220) | 2.6 (9) |
|  | **No impact (%)** | **Major impact (%)** | **No impact (%)** | **Major impact (%)** |
| Impact of T21 on business operations | 23.5 (87) | 19.1 (88) | 30.0 (105) | 15.1 (52) |

*Note: Percentages of sample are weighted.*

Most retailers, pre and post (63%, 64%) felt that it would be/has been easy for their store to comply with the new sales law. Of those who said it would be hard to comply with the law, 36% in the pre-T21 survey believed that the law would make customers upset and over half (53%) post-T21 cited upset customers as the major difficulty of compliance. Difficulty disseminating and educating customers was a concern shared by 18% of retailers pre-T21 but decreased to five percent post-T21.

Retailers were asked a series of questions on both surveys to gauge their understanding of the law and overall perceptions of tobacco products and policy (Table 3). Data showed retailers had a good understanding of T21 and how to comply both pre-T21 and post-T21 (91% agreed) and afterward (93% agreed). More than two-thirds also said they knew where to get information about T21 compliance after the law went into effect (81% pre-T21, 89% post-T21). In both survey waves, over 70% of retailers agreed that increasing the sales age to 21 makes it harder for those under 21 to obtain tobacco products.

**Table 3:**
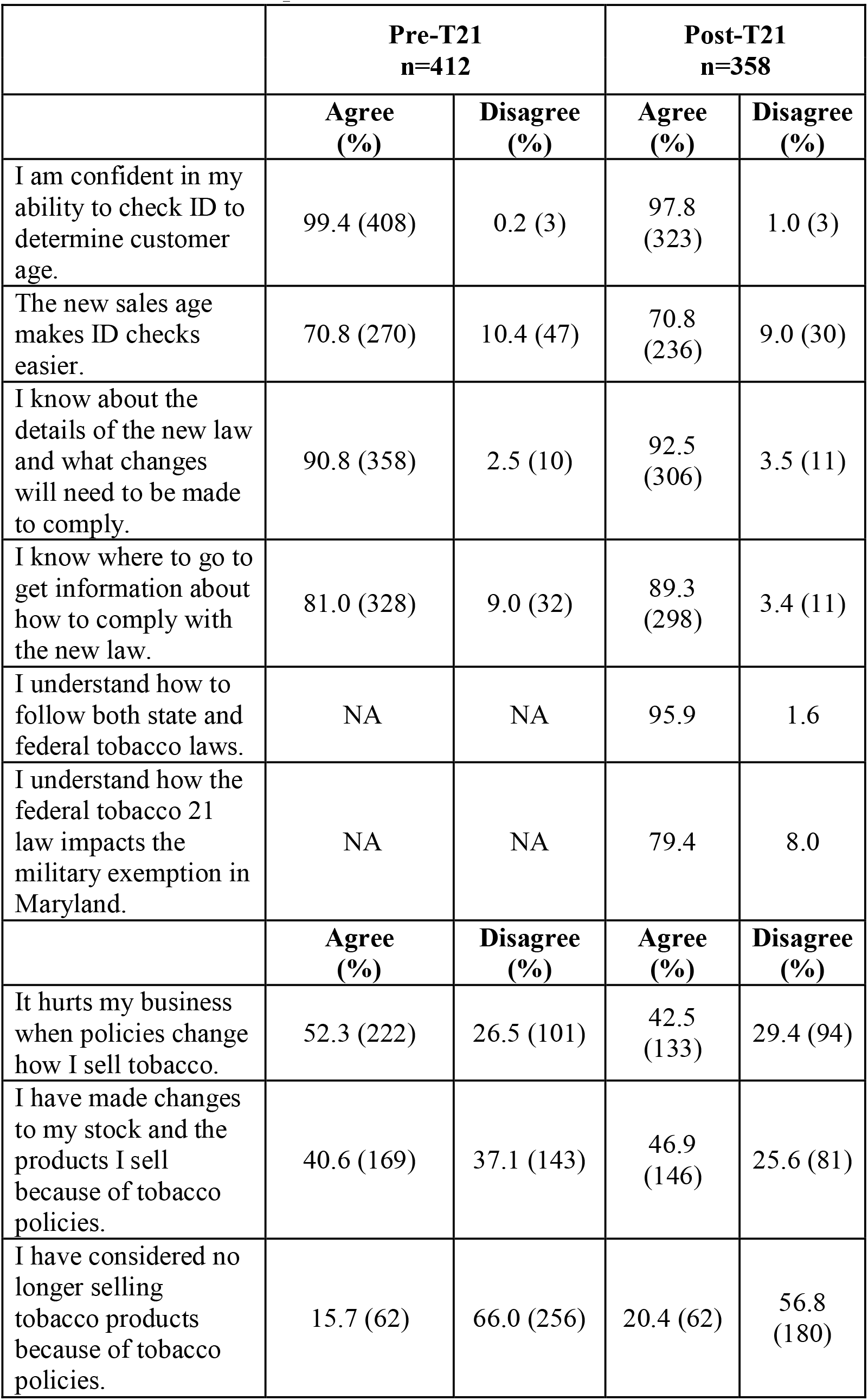
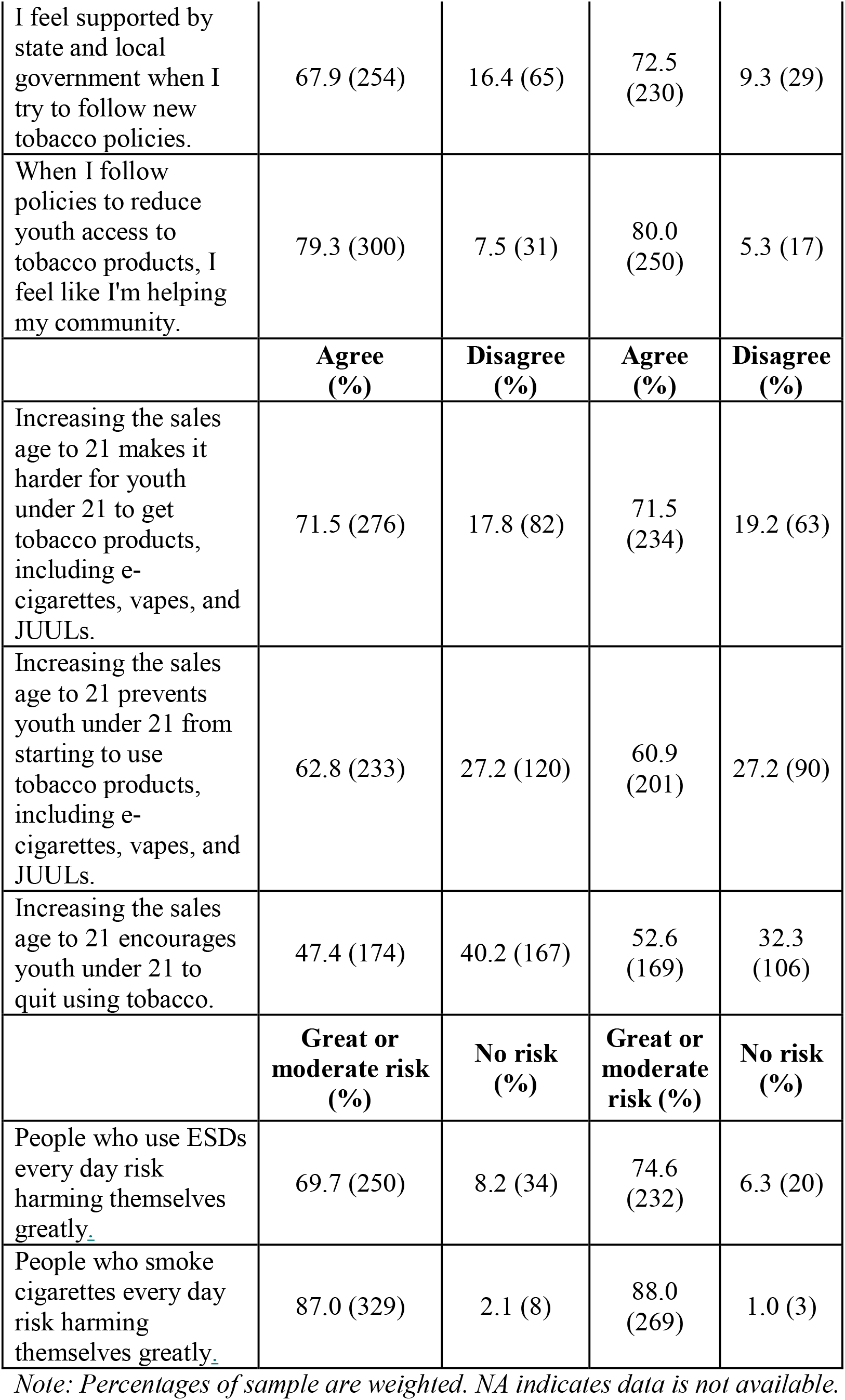
Retailer Perceptions.

The federal law went into effect on December 20, 2019 and had the potential to cause confusion regarding the difference in state and federal legislation. However, most retailers were confident that they understood how to follow both state and federal tobacco control laws. Maryland’s T21 law contained a military exemption that would allow active duty military members with ID to purchase tobacco. There is no such exemption in the federal law, and this discrepancy was noted by a small number of retailers who reported the difference made it challenging to comply with the correct law, despite MDH providing guidance noting that federal law supersedes state law.

Retailers were also asked their opinions about how harmful it is to smoke cigarettes or use ESDs. More than half (88%) in the post-survey felt smoking cigarettes every day was a moderate to great risk, compared to 75% who felt the same about ESDs.

As shown in Table 4, among youth who attempt to purchase tobacco products, cigarettes are the most common product attempted to purchase, followed by cigars, blunt wraps, and ESDs. Over a quarter of retailers strongly agree that customers under 21 prefer ESDs over traditional tobacco products.

**Table 4:** Tobacco Customer Preferences.

|  | **Pre-T21**<br>**n=403**<br>**%** | **Post-T21**<br>**n=325**<br>**%** |
| --- | --- | --- |
| *Tobacco products youth usually ask to buy at store* |  |  |
| Cigarettes | 56.3 (222) | 47.3 (154) |
| Cigars, little cigars, cigarillos | 50.9 (206) | 43.7 (143) |
| Blunt wraps or rolling papers | 30.9 (125) | 39.5 (129) |
| ESDs | 28.9 (129) | 31.0 (103) |
| Smokeless tobacco | 10.5 (42) | 10.9 (36) |
| None of the above | 8.0 (30) | 17.7 (58) |
| Customers under 21 prefer flavored tobacco products and ESDs (e-cigarettes, vapes, JUULs). (% Strongly Agree) | 32.5 (123) | 29.1 (93) |
| Customers under 21 prefer ESDs (e-cigarettes, vapes, JUULs) to traditional tobacco products (cigarettes, cigars, etc.). (% Strongly Agree) | 27.2 (101) | 27.1 (86) |
| ESDs (including e-cigarettes, vape pens, pod devices such as Juul) should only be sold in adult-only stores. (% Strongly Agree) | 35.3 (141) | 34.1 (108) |

## Discussion

The purpose of this study was to measure the level of retailer awareness, support, and challenges to comply with the new Tobacco 21 law, as well as identify opportunities to better support and educate retailers regarding T21, to reduce youth access to tobacco products in the retail environment.

This study is among the first of its kind to evaluate the perception and preparedness for T21 before and after the law passage, in order to gauge compliance, opinions and challenges related to T21’s success.

The first wave of this survey sought out retailers’ opinions in Maryland immediately before the T21 law went into effect. About one-fifth of retailers voiced concerns about how the law would impact revenue, but post T21 results show that by and large retailers felt they had not been impacted financially and that the law only had minor effects on their business operations. Of note, considerably more vape shops said the law had a significant impact on their business operations. This implies that a higher portion of vape shop sales were made to customers under 21 years old prior to T21. This also indicates that youth are frequently purchasing ESD products from these retailers, a concern since vaping rates have increased significantly in the past five years. While youth ESD prevalence rates are not yet available for Maryland post T21, a key indicator going forward will be the impact of T21 on smoking and ESD prevalence among youth.

Overall, retailers thought it would be easy to comply with T21 and that was shown to be true after implementation. Nearly all retailers were confident in their ability to check ID and their basic understanding of the law. Support of the law remained consistent both pre and post implementation, which may reflect that the law did not significantly impact their business. The relative ease of T21 compliance at the retail level may also be related to the concerted retailer outreach efforts conducted by MDH before the law went into effect.

The study found that retailers were satisfied with the resources and support they received from MDH. Most were aware of how to obtain information and felt confident in their ability to comply with the law. Awareness was also high regarding the free “21 or None” materials, including an ID guide, posters, window adhesives and quick reference guide, sent to retailers by MDH and most retailers found them useful. MDH took several steps through the summer and fall of 2019 to increase awareness about the law to both retailers and residents and this effort appears to have been effective in communicating the T21 changes to retailers and helping them understand what to expect.

MDH spent months distributing educational and promotional materials to retailers prior to the implementation of T21, including information about the active-duty military exemption, as well as window clings and posters to promote the law within the store. Retailers believed that there was a significant amount of education geared toward retailers, but many noted it would be helpful to have additional advertising directed towards increasing public awareness of the law. Given that retailers do not consider it their role to educate consumers, retailer feedback suggests that educating the public at large would decrease, over time, the negative feelings associated with the change in the law and decrease the strain put on retailers to educate the public about the new law while trying to maintain their business and keep their employees safe.

The law did not appear to significantly impact retailer ID check or carding practices. While two-thirds of retailers said they card or check IDs more often because of T21, how they conduct their checks did not drastically change from the pre to post-T21 survey. Most retailers continued to card those that look under a certain age rather than carding all customers as a rule.

Maryland’s T21 law included a state-level exemption for military personnel with an active-duty military ID. A few states, California among them, had a T21 law prior to the federal implementation that also included a military exemption.^xiii^ When retailers were asked their opinion about the military exemption, many had concerns. Among those, 16% thought the exemption should apply to all military personnel (vs just active duty), while 13% said the sales age should be 21 years old for everyone. When the federal law was enacted in late December 2019 with no federal exemption, this preempted and, in effect, removed the military exemption in Maryland’s state law.

While many retailers stated that members of the military under age 21 should be able to purchase tobacco, studies show the use of tobacco products adversely impacts military readiness. The CDC published a report stating T21 laws without a military exemption have the added benefit of not only promoting public health, but also national security goals. The surgeons general of the Air Force, Army, Navy, and United States, as written in a joint letter to the Department of Defense, strongly recommended that tobacco laws not include a military exemption. ^xiv^

Despite the change in laws and policies, most retailers are not inclined to stop selling tobacco products. As shown in this study and others, most people have adapted to the changing laws and agree that increasing the sales age for tobacco products is a positive thing. That T21 is now a federal law shows the increased commitment to reduce tobacco access and, ultimately, decrease the number of youth tobacco users.

The evaluation of T21 is an important tool in determining both the law’s success and the reasons it succeeded or did not. The risk of noncompliance increases if retailers either do not understand or disagree with the law. This study not only asked retailers how the law impacted them after implementation, but also evaluated how they thought it might impact their business and practices prior to the enactment of T21. Retailers were able to share their opinions, worries and perceived impact of the law and then MDH was able to follow up and see if these concerns came to fruition. This helps to identify gaps in understanding and help state and federal agencies evaluate their efforts and improve efficacy. As this study demonstrates, retailers had few complaints related to adherence and, overall, had a positive impression of the promotion and training efforts prior to T21.

### Limitations

Federal Tobacco 21 legislation went into effect on December 20, 2019, roughly two months after Maryland’s T21 went into effect. While similar in many ways, the military exemption was a key difference. Per federal law, military personnel have the same restricted access to tobacco and related products as non-military residents. The state of Maryland had spent the months between passing legislation and the implementation date of T21 educating retailers of the laws and how to comply, advising them of the military exemption for active duty military between 18 and 20 years old. This caused confusion when only a couple months later federal law effectively removed this exemption. Retailers noted this confusion, and, for some, it was unclear if federal law superseded the state law, although MDH released guidelines stating that federal law would take precedence.

The tobacco retailer market changed between the pre-T21 and post-T21 surveys as the United States entered a period of social distancing and quarantine due to COVID-19. In Maryland, convenience stores and gas stations were deemed essential and were able to remain open to the general public. Vape shops and tobacconists, on the other hand, were not deemed an essential business and were closed from March 23, 2020 to May 15, 2020.^xv xvi^ Vape shop retailers accounted for seven percent of the respondents during the pre-T21 survey but dropped to one percent during the post-T21 survey. While this may be linked to the closures and retailers not being accessible, the rates for tobacconist retailers remained consistent, at five percent of respondents’ pre and post, despite being under the same orders.

Beyond COVID-19 closures, the respondents’ investment in the issue and concerns about compliance and awareness may have decreased during the pandemic. Issues that had been of high importance garnered less attention as more immediate concerns took precedence. Retailers were likely dealing with decreased revenue, keeping workers safe, complying with ever-changing protocols and fear of contracting or spreading the virus.

This research relied on self-reported data, in and of itself a limitation. Respondents hold a personal bias that influences how they perceive and interpret information. Self-reported answers may be exaggerated and, information may be withheld due to social desirability bias.

## Data Availability

Data is available upon reasonable request

## Appendix Questionnaire

### Tobacco Retailer Survey

Center for Tobacco Prevention and Control, Maryland Department of Health August 20, 2019

### Introduction

The Maryland Department of Health is conducting an important survey of tobacco retailers and vape shops across the state. **The Department would like to hear your opinion on how laws that regulate tobacco sales affect your store and how best to support your business to ensure compliance with these laws.**

The questions take about 10 minutes to answer. Your participation is voluntary. We do not ask your name or the name of your store.

Your responses are kept confidential and will only be reported in combined form with the responses of others. The survey is being conducted by a research firm outside of Maryland. Your responses will **not** impact retail enforcement of your business.

Each store should answer the survey only one time. The person who answers the survey should be someone who knows about the policies and procedures that take place in the store.

The survey is run by Market Decisions Research, an independent public health evaluation firm. If you have any questions about the survey, please contact the study director, Patrick Madden of Market Decisions Research at, or call toll-free at 1-800-293-1538, extension 101.

The Maryland Department of Health’s Institutional Review Board has reviewed the survey. If you have questions about the protection of survey participants, contact Gay Hutchen, Chief Administrator, IRB and Liaison Unit, Maryland Department of Health at *, or 410-767-8448*. Questions about tobacco policies can be submitted to the Maryland Department of Health at.

Please click on the button below to begin the survey.

#### Screening Questions

1. Does your store sell tobacco products, such as cigarettes, cigars or cigarillos, smokeless tobacco, or electronic smoking devices, such as e-cigarettes, vapes, pod devices such as JUUL®, and e-liquids?
  • Yes
  • No [TERMINATE]

#### Awareness and Perception of Law

2. Are you aware of a new Maryland law that raises the minimum sales age from 18 to 21 for all tobacco products (including all electronic smoking devices such as e-cigarettes, vapes, pod devices such as JUUL®, and e-liquids)?
  • Yes
  • No
  • Not Sure
3. Do you support the new law that raises the minimum sales age for tobacco from 18 to 21?
  1. Support
  2. Somewhat support
  3. No opinion
  4. Do not support
4. Are you aware that the new law allows retailers to sell tobacco to active duty military under 21 if they have valid military ID?
  • Yes
  • No
  • Not Sure
5. What are your thoughts on the exemption in the law that allows retailers to sell tobacco to active duty military under 21?
  1. The law should not allow sales to anyone under 21
  2. I agree with the exemption as written
  3. No opinion
6. How much of an impact will the new sales law have on your business operations?
  1. Major impact
  2. Minor impact
  3. None
7. How easy will it be for your store to comply with the new sales law when it goes into effect?
  1. Easy
  2. Somewhat Easy
  3. Somewhat Difficult
  4. Difficult
8. [ASK IF SOMEWHAT DIFFICULT OR DIFFICULT IN Q7] Why will it be difficult to comply with the new sales law? (Choose the top reason)
  10. It costs too much money to make changes
  11. It is difficult to notify customers
  12. It makes customers upset
  13. It hurts my business
  14. It’s too hard to re-do my displays
  15. I need more information about policies
  16. It’s difficult to train employees to follow new policies
  17. It is too complicated to check ID
  18. It is too complicated to check military IDs
  19. Other (Specify)
  20. None of the above
9. Do you agree or disagree with the following statements? I am confident in my ability to check ID to determine customer age. The new sales age makes ID checks easier. I know about the details of the new law and what changes will need to be made to comply. I know where to go to get information about how to comply with the new law. Increasing the sales age to 21 makes it harder for youth under 21 to get tobacco products, including e-cigarettes, vapes, and JUULs Increasing the sales age to 21 prevents youth under 21 from starting to use tobacco products, including e-cigarettes, vapes, and JUULS. Increasing the sales age to 21 encourage youth under 21 to quit using tobacco.
  • Strongly agree / Somewhat agree / Neither / Somewhat disagree / Strongly disagree
10. Will the state law that increases the minimum sales age for tobacco from 18 to 21 years old affect how often you card or check IDs of customers trying to buy tobacco?
  1. Yes, I plan to card or check IDs **more**
  2. Yes, I plan to card or check IDs **less**
  3. No, I will not change my carding practices
  4. Don’t know
11. What additional information or resources would make it easier for you to comply with the law that raises the minimum age to sell tobacco in the state to 21? *[open-ended]*

#### Store Policies and Products

12. How do you or your employees decide which customers to “card” or ask for identification when buying tobacco? *[open-ended]*
13. Among your usual customers who buy tobacco products, how many are under 21 years of age?
  • Most
  • Some
  • A Few
  • None
14. Which tobacco products do customers under 21 usually buy at your store? Select all that apply.
  1. Cigarettes
  2. Cigars, Little Cigars, Cigarillos
  3. Blunt wraps or rolling papers
  4. Electronic smoking devices (electronic cigarettes, vape pens, pod devices such as JUUL®) or e-liquids
  5. Smokeless tobacco (“snus”, “snuff”, “chew”, “dip”)
15. Do you agree or disagree with the following statements? Customers under 21 prefer flavored tobacco products and electronic smoking devices (e-cigarettes, vapes, JUULs). Customers under 21 prefer electronic smoking devices (e-cigarettes, vapes, JUULs) to traditional tobacco products (cigarettes, cigars, etc.). Electronic smoking devices (including electronic cigarettes, vape pens, pod devices such as JUUL^®^) should **only** be sold in adult-only stores.
  • Strongly agree / Somewhat agree / Neither / Somewhat disagree / Strongly disagree
16. How frequently do you sell tobacco products to customers with military ID?
  1. Every day
  2. Most days
  3. Occasionally
  4. Rarely
  5. Never
17. The next few questions ask about your familiarity with the exemption for military in the new sales age law.
  • I am familiar with military ID.
  • I know how to check military ID for **active** duty status.
  • I am comfortable refusing sales to military if they do not qualify for the exemption.
  • It is easier to have the same minimum sales age for all customers.
  • Yes/No
18. Do you agree or disagree with the following statements? It hurts my business when policies change how I sell tobacco. I have made changes to my stock and the products I sell because of tobacco policies. I have considered no longer selling tobacco products because of tobacco policies. I feel supported by state and local government when I try to follow new tobacco policies. When I follow policies to reduce youth access to tobacco products, I feel like I’m helping my community.
  • Agree / Somewhat agree / Neither / Somewhat disagree / Disagree

#### Retailer Perception and Attitudes toward Tobacco and Health

19. In your opinion, how much do people who use electronic smoking devices (e-cigarettes, vapes, JUUL®) everyday risk harming themselves?
  • No risk
  • Slight risk
  • Moderate risk
  • Great risk
20. In your opinion, how much do people who smoke (cigarettes, cigars) every day risk harming themselves?
  • No risk
  • Slight risk
  • Moderate risk
  • Great risk

#### About You

21. What is your role in the store? Are you the:
  • Owner
  • Manager
  • Clerk
  • Other (Specify)
22. Which type of store best describes your store?
  10. Convenience store (without gas)
  11. Convenience store (with gas)
  12. Pharmacy
  13. Liquor store
  14. Grocery store
  15. Mass merchandisers
  16. Tobacco shop
  17. Vape shop
  18. Other (Specify)
23. [ASK OF VAPE SHOPS ONLY FROM Q22] Do you currently sell electronic smoking devices or accessories online?
  • Yes
  • No
24. [ASK IF YES TO 23] How do you verify customer age?
  1. A pop-up box that asks for date of birth or for user to click “Yes, I’m of legal age.”
  2. Third-party age verification software
  3. Don’t check customer age
  4. Other (Specify)
  5. Don’t know
25. If you would be interested in participating in a similar survey after the new minimum sales age law goes into effect, please enter your email address below. Please note that your email will remain confidential, will only be used for the purposes of follow-up for this survey, and it will not be stored or associated with the answers that you provided here.

**Thank you for your time. That completes the survey!**

**If you have any questions about the new minimum sales age for tobacco products, please visit www.notobaccosalestominors.com or contact the Maryland Department of Health at.**

### Tobacco Retailer Post Survey

Center for Tobacco Prevention and Control, Maryland Department of Health

April 2020

#### Introduction

The Maryland Department of Health is conducting an important survey of tobacco retailers and vape shops across the state. **The Department would like to hear your opinion on how laws that regulate tobacco sales affect your store and how best to support your business to ensure compliance with these laws.**

The questions take about 10 minutes to answer. Your participation is voluntary. We do not ask your name or the name of your store.

Your responses are kept confidential and will only be reported in combined form with the responses of others. The survey is being conducted by a research firm outside of Maryland. Your responses will not impact retail enforcement of your business.

Each store should answer the survey only one time. The person who answers the survey should be someone who knows about the policies and procedures that take place in the store.

The survey is run by Market Decisions Research, an independent public health evaluation firm. If you have any questions about the survey, please contact the study director, Patrick Madden of Market Decisions Research at, or call toll-free at 1-800-293-1538, extension 101.

The Maryland Department of Health’s Institutional Review Board has reviewed the survey. If you have questions about the protection of survey participants, contact Gay Hutchen, Chief Administrator, IRB and Liaison Unit, Maryland Department of Health at *, or 410-767-8448*. Questions about tobacco policies can be submitted to the Maryland Department of Health at.

Please click on the button below to begin the survey.

#### Screening Questions

26. Does your store sell tobacco products, such as cigarettes, cigars or cigarillos, smokeless tobacco, or electronic smoking devices, such as e-cigarettes, vapes, pod devices such as JUUL®, and e-liquids?
  • Yes
  No [TERMINATE]

#### Awareness and Perception of Law

27. Are you aware of a new Maryland law that raises the minimum sales age from 18 to 21 for all tobacco products (including all electronic smoking devices such as e-cigarettes, vapes, pod devices such as JUUL®, and e-liquids)?
  • Yes
  • No
  • Not Sure
28. Are you aware of a new Federal law that raises the minimum sales age from 18 to 21 for all tobacco products (including all electronic smoking devices such as e-cigarettes, vapes, pod devices such as JUUL®, and e-liquids)?
  • Yes
  • No
  • Not Sure
  *Beginning October 1 2019, Maryland’s ‘Tobacco 21’ law went into effect, raising the minimum legal sales age of tobacco products from 18 to 21 years old. At the end of 2019, the United States federal government raised the minimum tobacco sales age from 18 to 21 years old across the country.*
29. Do you support the new law that raises the minimum sales age for tobacco from 18 to 21?
  • Support
  • Somewhat support
  • No opinion
  • Do not support
30. How much of an impact has the new sales law had on your business operations?
  • Major impact
  • Minor impact
  • None
31. Are you currently selling tobacco products to active duty military under 21?
  • Yes
  • No
  • Not Sure
32. What were your thoughts on the exemption in the state law that allowed retailers to sell tobacco to active duty military under 21? *[open-ended]*
33. How easy has it been for your store to comply with the new sales law since it has gone into effect?
  • Easy
  • Somewhat Easy
  • Somewhat Difficult
  • Difficult
34. [ASK IF SOMEWHAT DIFFICULT OR DIFFICULT IN Q7] Why has it been difficult to comply with the new sales law? (Choose the top reason)
  • It costs too much money to make changes
  • It is difficult to notify customers
  • It makes customers upset
  • It hurts my business
  • It’s too hard to re-do my displays
  • I need more information about policies
  • It’s difficult to train employees to follow new policies
  • It is too complicated to check ID
  • Confusion between state and federal laws
  • Confusion about the military exemption
  • Underage youth do not know the law
  • Other (Specify)
  • None of the above
35. Do you agree or disagree with the following statements?
  a. I am confident in my ability to check ID to determine customer age.
  b. The new sales age makes ID checks easier.
  c. I know about the details of the new law and what changes will need to be made to comply.
  d. I know where to go to get information about how to comply with the new law.
  e. Increasing the sales age to 21 makes it harder for youth under 21 to get tobacco products, including e-cigarettes, vapes, and JUULs.
  f. Increasing the sales age to 21 prevents youth under 21 from starting to use tobacco products, including e-cigarettes, vapes, and JUULS.
  g. Increasing the sales age to 21 encourages youth under 21 to quit using tobacco.
  h. I understand how to follow both state and federal tobacco laws.
  i. I understand how the federal tobacco 21 law impacts the military exemption in Maryland.
    • Strongly agree / Somewhat agree / Neither / Somewhat disagree / Strongly disagree
36. Did the state law that increases the minimum sales age for tobacco from 18 to 21 years old affect how often you card or check IDs of customers trying to buy tobacco?
  • Yes, I card or check IDs **more**
  • Yes, I card or check IDs **less**
  • No, I did not change my carding practices
  • Don’t know
37. Are you aware of the free “21 or None” materials from the Maryland Department of Health for retailers (such as signs, posters, window clings, etc.)?
  • Yes
  No
  Not Sure
38. Which of the resources provided by the Maryland Department of Health have been most useful to you? (select all that apply)
  • Age of Sale Poster
  • Age of Sale Window Adhesive
  • Quick Reference Guide
  • Product Identification Guide
  • ID Guide
  • Maryland and Federal Laws Chart
  • Local Laws Chart
  • Comprehensive Law Guide
  • Table Tent
  • Online Training
  • Training Quiz
  • Don’t know
  • Not applicable
39. What additional information or resources would make it easier for you to comply with the law that raises the minimum age to sell tobacco in the state to 21? *[open-ended]*

#### Store Policies and Products

40. How do you or your employees decide which customers to “card” or ask for identification when buying tobacco? *[open-ended]*
41. Among your usual customers, how many are youth under 21 years of age? If you aren’t sure, just take your best guess.
  • Most
  • Some
  • A Few
  • None
42. Which tobacco products do youth usually ask to buy at your store? Although these products are age-restricted by law, young people may still ask to purchase them. **Please select all the products that young people might ask to buy**.
  • Cigarettes
  • Cigars, Little Cigars, Cigarillos
  • Blunt wraps or rolling papers
  • Electronic smoking devices (electronic cigarettes, vape pens, pod devices such as JUUL®) or e-liquids
  • Smokeless Tobacco (“snus”, “snuff”, “chew”, “dip”)
  • None of the above
43. Do you agree or disagree with the following statements?
  a. Customers under 21 prefer flavored tobacco products and electronic smoking devices (e-cigarettes, vapes, JUULs).
  b. Customers under 21 prefer electronic smoking devices (e-cigarettes, vapes, JUULs) to traditional tobacco products (cigarettes, cigars, etc.).
  c. Electronic smoking devices (including electronic cigarettes, vape pens, pod devices such as JUUL®) should **only** be sold in adult-only stores.
    • Strongly agree / Somewhat agree / Neither / Somewhat disagree / Strongly disagree
44. At the beginning of 2020, how much of your total monthly sales were from <u>mint or menthol tobacco products</u>? (e.g. Newport Green cigarettes, JUUL Mint Pods)
  • None
  • 1 - 5%
  • 6 - 10%
  • 11 - 25%
  • 26 - 50%
  • 51 - 75%
  • 76 – 100%
45. At the beginning of 2020, had total monthly sales from <u>mint or menthol tobacco products changed from the previous year</u>?
  • Monthly sales had **increased**
  • Monthly sales had **decreased**
  • Stayed about the same
  • Not sure
46. How frequently do you sell tobacco products to customers with military ID?
  • Every day
  • Most days
  • Occasionally
  • Rarely
  • Never
47. Do you agree or disagree with the following statements?
  a. It hurts my business when policies change how I sell tobacco.
  b. I have made changes to my stock and the products I sell because of tobacco policies.
  c. I have considered no longer selling tobacco products because of tobacco policies.
  d. I feel supported by state and local government when I try to follow new tobacco policies
  e. When I follow policies to reduce youth access to tobacco products, I feel like I’m helping my community.
    • Agree / Somewhat agree / Neither / Somewhat disagree / Disagree

#### Retailer Perception and Attitudes toward Tobacco and Health

48. In your opinion, how much do people who use electronic smoking devices (e-cigarettes, vapes, JUUL®) everyday risk harming themselves?
  • No risk
  • Slight risk
  • Moderate risk
  • Great risk
49. In your opinion, how much do people who smoke (cigarettes, cigars) every day risk harming themselves?
  • No risk
  • Slight risk
  • Moderate risk
  • Great risk

#### New Policies

The following tobacco policies have been passed or have recently gone into effect in Maryland:

• A federal ban on the sale of <u>flavored, cartridge-based, electronic smoking devices</u>, including fruit, candy, and other non-tobacco or menthol flavors. Note, this <u>excludes</u> cigarettes, cigars or cigarillos, smokeless tobacco other non-electronic forms of tobacco, and non-cartridge-based electronic smoking devices.
• An increased enforcement of cartridge-based and disposable flavored electronic smoking devices by the Comptroller of Maryland, including additional license checks and product inspections to ensure retailer compliance.
• An increase in the state’s tobacco tax on electronic smoking devices and other tobacco products.

50. How much of an impact do these tobacco policies have on your business operations?
  • The federal ban on the sale of <u>flavored, cartridge-based, electronic smoking devices</u>
  • Increased enforcement of flavored electronic smoking devices
  • A tobacco tax Increase
    ∘ Major impact
    ∘ Minor impact
    ∘ None
51. How easy is it (or will it be) for your store to comply with the following policies?
  • The federal ban on the sale of <u>flavored, cartridge-based, electronic smoking devices</u>.
  • Increased enforcement of flavored electronic smoking devices.
  • A tobacco tax increase.
    ∘ Easy
    ∘ Somewhat Easy
    ∘ Somewhat Difficult
    ∘ Difficult
52. What method is best for the State to communicate to you about tobacco retail policies?
  • Standard Mail
  • Email
  • Post on NoTobaccoSalestoMinors.com website
  • ListServ
  • Social Media
  • Other (Specify)
53. In what language would you prefer to receive information about tobacco policies (Select all that apply)?
  • Arabic
  • Chinese, including Mandarin or Cantonese
  • English
  • French or French Creole
  • Hindi
  • Portuguese
  • Spanish
  • Urdu
  • Something else (Please tell us)

#### About You

54. What is your role in the store? Are you the:
  • Owner
  • Manager
  • Clerk
  • Other (Specify)
55. Which type of store best describes your store?
  • Convenience store (without gas)
  • Convenience store (with gas)
  • Pharmacy
  • Liquor store
  • Grocery store
  • Mass merchandisers
  • Tobacco shop
  • Vape shop
  • Other (Specify)
56. Which of the following tobacco products do you sell in your store or online? (select all that apply)
  • Cigarettes
  • Cigars, Little Cigars, Cigarillos
  • Menthol flavored tobacco products
  • Flavored cigars/cigarillos/little cigars
  • Blunt wraps or rolling papers
  • Electronic cigarettes (“e-cigarettes”), vape pens, other electronic vaping devices, or e-liquids
  • Juul device/Juul pods
  • Puff bars
  • Suorin products
  • Flavored e-juice, including pods
  • Smokeless Tobacco (“snus”, “snuff”, “chew”, “dip”)
  • Other (Specify)
  • None of the above

**Thank you for your time. That completes the survey!**

**If you have any questions about the new minimum sales age for tobacco products, please visit www.notobaccosalestominors.com or contact the Maryland Department of Health at.**

## Notes

### Competing Interest Statement

The authors have declared no competing interest.

### Funding Statement

Authors are funded through Maryland Department of Health.

### Author Declarations

Maryland Department of Health Institutional Review Board

